# Validation of the MAGIC genomic signature on RNAseq in soft-tissue sarcomas

**DOI:** 10.1101/2025.07.19.25331113

**Authors:** Ataaillah Benhaddou, Gaëlle Pérot, Philippe Rochaix, Thibaud Valentin, Gwenaël Ferron, Frédéric Chibon

## Abstract

**Background:** Genomic Instability (GIN) plays a critical role in cancer progression and treatment response. Soft tissue sarcomas (STS), are characterized by high levels of chromosomal rearrangements and transcription-associated stress, both of which contribute to poor clinical outcomes. Current standard grading systems, such as FNCLCC, are limited in prognostic accuracy for STS, necessitating novel approaches for risk stratification and treatment guidance. To address this gap, we developed a holistic classifier, the MAGIC (Mixed transcription- and replication-associated GIN classifier), combining transcription- and replication-related GIN indices (iTRAC and iRACIN) to predict metastatic risk.

**Patients and Methods:** This study utilized RNA sequencing (RNAseq) on 226 STS tumor samples to analyze fusions transcript’s break points (BP) distribution and assess GIN. We computed MAGIC indices iTRAC and iRACIN, which are based on chromosomal instability linked to transcription and replication processes, respectively. iTRAC biomarker was evaluated against FNCLCC and CINSARC for metastatic risk stratification. Kaplan-Meier and iPART analyses were used to determine the prognostic relevance of iTRAC levels.

**Results:** iTRAC significantly stratified patients with distinct metastatic outcomes, outperforming FNCLCC and CINSARC grading systems. STS patients with medium level of iTRAC showed the poorest metastasis-free survival. Patients classified as iTRAC high- and low-risk groups, achieved better prognosis. Furthermore, iTRAC stratified patients’ metastatic risk in treated and not treated patients, indicating poorer prognosis with chemotherapy in patients with low iTRAC and better for those with with medium iTRAC.

**Conclusion(s):** iTRAC demonstrates a superior prognostic utility in STS over current grading systems, effectively stratifying metastatic risk for patients who might benefit from alternative therapeutic strategies. iTRAC holds potential for personalizing chemotherapeutic approaches, paving the way for a new precision oncology approach in STS.

## Introduction

The non-linear relationship between GIN and clinical outcome is a recent and paradigm shifting concept in genomics and cancer therapeutics (Birkbak *et al*. 2011; Roylance *et al*. 2011; Andor *et al*. 2016; Benhaddou *et al*. 2021;). Several cancers were reported to show a non-linear relationship between GIN and clinical outcome comprising breast, ovarian, gastric, non–small cell lung cancers and leiomyosarcomas (Birkbak *et al*. 2011; Roylance *et al*. 2011; Benhaddou *et al*. 2021). This may have important implications for patient’s risk stratification and therapeutic choice. Interestingly, this non-linear relationship between GIN and prognosis has been proposed as the underlying mechanism why some gene expression approaches which are also correlated with GIN, such as Oncotype DX and MammaPrint, have limited prognostic value in ER-negative breast cancer (Roylance *et al*. 2011; Birkbak *et al*. 2011). Development of diagnostic methods that take into account this non-linear relationship between GIN and clinical outcome is therefore of paramount importance to leverage more efficiently cancer genomics for clinical care.

GIN is associated with poor prognosis in solid tumors (Carter *et al*. 2006; Walther, Houlston and Tomlinson 2008; Chibon *et al*. 2010) and the rate of aneuploidy and karyotypic abnormalities increases with tumor grade and invasiveness (Walther, Houlston, and Tomlinson 2008; Yamamoto *et al*. 2007; Florl and Schulz 2008; M’kacher *et al*. 2010; Florl and Schulz 2008). Conversely, experimental data show that chromosome missegregation and aneuploidy can also inhibit tumorigenesis (Weaver and Cleveland 2008). This is because aneuploidy may also cause proliferative disadvantage (Williams *et al*. 2008). Consequently, aneuploidy may have both tumor promoting and tumor suppressive effects in animal model systems (Schvartzman, Sotillo and Benezra 2010; Baker *et al*. 2009). Such a paradoxical relationship between GIN and clinical outcome suggests that there may have an optimal level of GIN for tumor progression, beyond which further instability generates non-viable karyotypes and provides no growth advantage that may even be deleterious for cancer cell fitness and aggressiveness (Cahill *et al*. 1999). Computational work corroborates this idea and predicts that a chromosome missegregation rate between 10^−3^ and 10^−2^ per chromosome is optimal for cellular fitness (Komarova and Wodarz 2004). The same idea was proposed for mutation rates for the fitness of population in dynamic environment (Nilsson *et al*. 2002).

MAGIC is a genomic signature, composed of two biomarkers and an algorithm which explores the relationship between GIN and clinical outcomes, which was first developed on whole genome sequencing (WGS) of leiomyosarcomas (Benhaddou *et al*. 2021). MAGIC employs a random breakage model to assess the enrichment of BP in selected structural and/or functional genomic sequences. It consists of two biomarkers that measure the concentration of DNA BP associated with transcription (iTRAC) and replication (iRACIN). Using MAGIC approach, we have shown that a medium level of iTRAC and iRACIN is associated with poor clinical outcome compared to low and high levels. We have also shown that in iTRAC-Low patients group, patients treated with chemotherapy have increased metastatic risk compared to not-treated patients. No statistically significant difference between chemotherapy treated and not treated patients has been observed in iTRAC-Medium and High groups.

MAGIC has implications in precision oncology but WGS is not currently the first choice in routine clinic. We therefore sought to translate MAGIC on RNAseq, a widely used technique in the clinics because it is cheaper and allows gene expression quantification for functional analysis of genes expression programs for multiple diagnostic and R&D purposes. To do so, we used the same cohort of leiomyosarcomas published in Benhaddou *et al*. 2021 and a second independent cohort previously published by our team (Lesluyes *et al*. 2016) of pleomorphic soft-tissue sarcomas for which RNAseq data were available. We combined the two cohorts into one single Pan-soft tissue sarcoma cohort (PanSTS) of 226 patients (Supplementary table 1) and reanalysed RNAseq data to detect BP.

## Methods

### RNAseq data

The RNA-seq data utilized in this study were sourced from studies previously published by our team: 1) Darbo *et al*. 2023 and 2) Lesluyes *et al*. 2016 which can be downloaded from the following links: https://ega-archive.org/studies/EGAS00001001262 and https://www.ncbi.nlm.nih.gov/geo/query/acc.cgi?acc=GSE71119 respectively.

### Bioinformatics analysis pipeline for RNA-seq

We estimated general read quality using FastQC (v0.10.1), trimmed low-quality 5’ and 3’ ends (Phred score <20) using TrimGalore (https://github.com/FelixKrueger/TrimGalore). Transcriptomic (coding RefSeq genes from Hg38 UCSC Table Browser (Kuhn, Haussler, et Kent 2013) fixed on 2021/03/18) alignments were performed by STAR (v2.6.0c) (Dobin *et al*. 2013). Once aligned, we removed paired-end reads that mapped at multiple locations on transcriptome, low-quality (score <20) and discordant paired-end reads using SAMtools (v0.1.19)(Li *et al.* 2009). MarkDuplicates (PicardTools v1.99 “Picard Toolkit.” 2018. Broad Institute, GitHub Repository. http://broadinstitute.github.io/picard/; Broad Institute) removed PCR duplicates and Cufflinks (v2.1.1) (Trapnell *et al*. 2012) estimated gene and transcript abundances. HTSeq (v0.5.4) (Anders, Pyl, et Huber 2015) numbered reads mapped to Spike-in and reference chromosomes. We used Arriba (Uhrig *et al*. 2021), for the detection of fusion transcripts from RNA-Seq data with defaults parameters.

### Statistical analysis

All statistical tests were carried out using R version 4.1.2 (2021-11-01);R: A language and environment for statistical computing. R Foundation for Statistical Computing, Vienna, Austria;http://www.r-project.org/index.html). Kaplan-Meier method was used for survival curves construction. Log-rank test was used to compare survival curves between groups. Cox regression was used for multivariate analysis.

### iPART, iTRAC, iRACIN, BP count

iPART is the stratification algorithm used to split cohort into groups of different levels of iTRAC and iRACIN and was applied as published in Benhaddou *et al*. 2021. iTRAC and iRACIN biomarkers were computed as described in Benhaddou *et al*. 2021. Total BP count for each patient was computed as the sum of split read coverage of all BP in each patient. Similarly, BP count for each iTRAC and iRACIN biomarkers was computed as the sum of split read coverage of each BP present in each biomarker. Here how it is done:

The readable genome is represented as a single interval of length L in base pairs (bp). The uniform probability Pu of any genomic position to carry a BP is computed as:

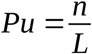

where:

- n is the total number of BP, now computed as the sum of split read coverages over all BP,
- L=2,948,611,470 bp is the total readable genome size (total ungapped genome length from NCBI assembly GCF_000001405.26).

For a given genomic interval of size Li and ni BP, the probability of harboring ni BP under the random breakage model (RBM) is computed using the binomial probability mass function:

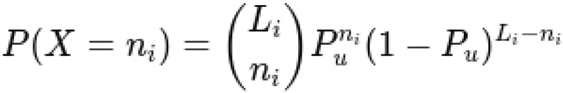

where:

- X is the random variable representing the number of BP in the interval,
- n_i_ is the **total BP count** in the interval (sum of split read coverages over all BP in the interval),
- L_i_ is the **size of the interval** (sum of merged interval sizes),
- P_u_ is the **uniform probability of BP occurrence**.

The probability of observing **more than n_i_ BP** under RBM is:

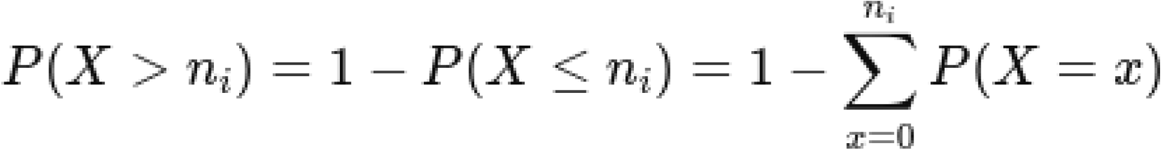

The **H-score** is then computed as:

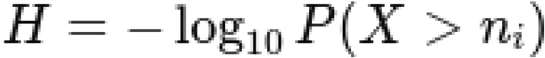

## Results

### Medium level of iTRAC measured on fusion transcripts BP is associated with poor clinical outcome

Using WGS we previously showed that medium levels of iTRAC and iRACIN are predictive of poor clinical outcome (Benhaddou *et al*. 2021). We therefore sought to check whether this observation still holds using RNAseq. We computed Hscore for iTRAC and iRACIN on the total of the fusion transcripts BP identified in the whole panSTS cohort (226 patients). We found that for iTRAC in the whole cohort, an Hscore equal to 350 (expected BP count = 21028, observed BP count =41148) meaning that fusion transcripts BP are highly overrepresented in iTRAC DNA elements compared to what is expected by chance. Fusion transcripts BP was depleted in iRACIN, with an Hscore equal to 0 (expected BP count =32654, observed BP count =3943). Indeed iRACIN DNA elements are more associated with replication related GIN. Therefore, it is only iTRAC that is relevant as a biomarker applied to the transcriptome of this data set. We then computed iTRAC for each individual patient and applied the iPART algorithm (Benhaddou *et al*. 2021) to split the patients of PanSTS cohort which did not receive chemotherapy (167 patients) into 3 groups of distinct levels of iTRAC: Low, Medium and High. Kaplan-Meier analysis shows that application of iTRAC over RNAseq fusion transcripts BP significantly stratified the untreated (*i.e*. without chemotherapy) panSTS cohort (P< 0.0001) into three groups with nonlinear relationship between iTRAC Hscore and clinical outcome (MFS) (fig 1A). Concordantly with results obtained with WGS, it is the group of patients with medium iTRAC that has the most unfavorable outcome compared to the reference group (Low iTRAC) with hazard ratio (HR) of 5 (P=1.57×10^-8^) (fig 1B). We also confirmed that Low and High iTRAC have similar clinical outcome (HR=1, P=4.28×10 ^-1^) (fig 1B).

**Figure 1:**
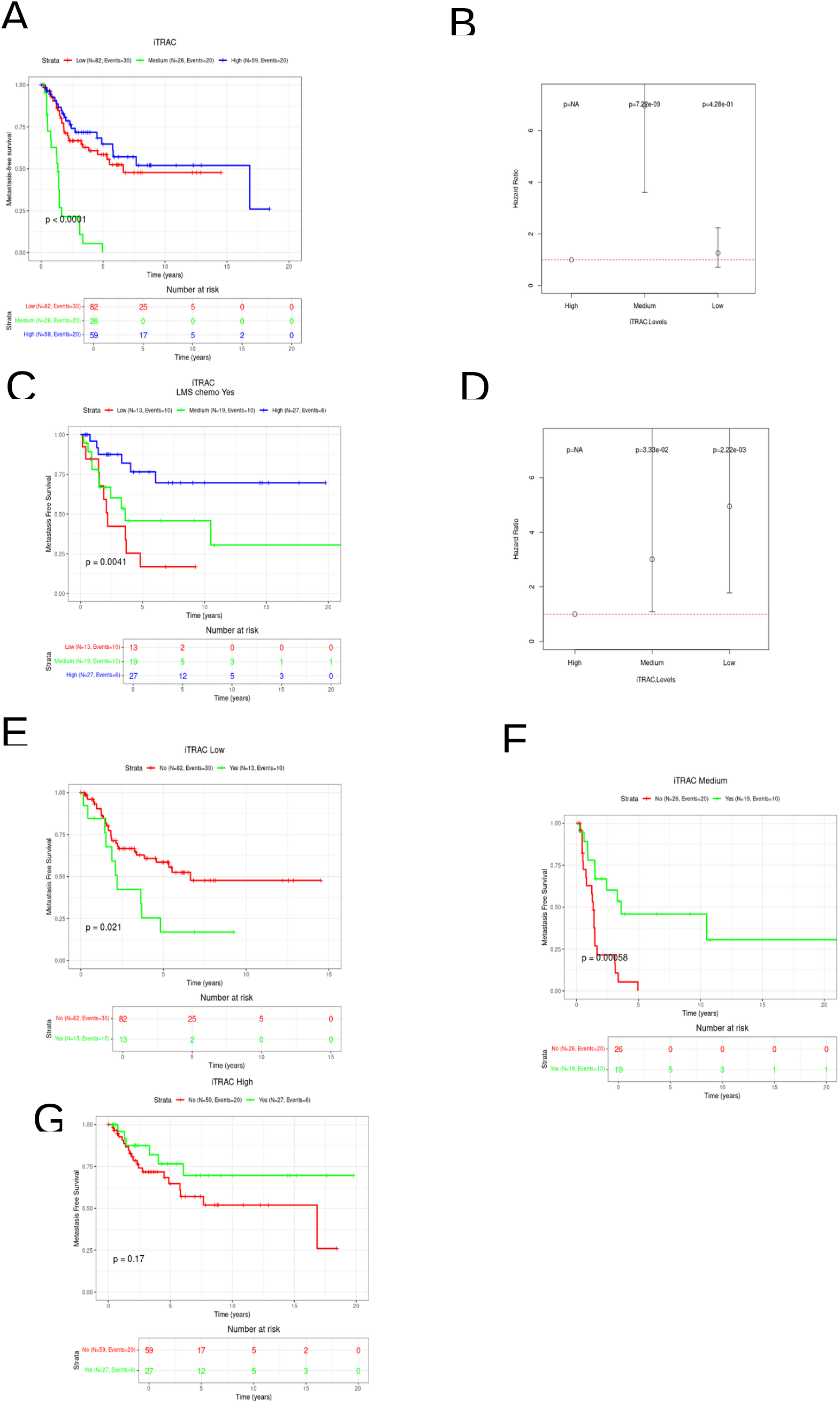
In each iTRAC stratification level, metastasis-free survival (MFS) outcomes differ between patients treated with chemotherapy and those untreated. A. Kaplan–Meier survival plot showing the panSTS cohort of iTRAC patients who were not treated with chemotherapy, stratified into Low, Medium, and High iTRAC levels. B. Hazard Ratio (HR) with a 95% confidence interval (CI) for the risk of metastasis in each ITRAC group relative to the Low group for the non-chemotherapy-treated panSTS cohort. P-values for each HR are displayed above each condition. The dashed horizontal line indicates an HR of 1. C, D, E. Kaplan–Meier plots for chemotherapy-treated and untreated patients in the Low, Medium, and High iTRAC groups, respectively. F. Kaplan–Meier plot of the chemotherapy-treated panSTS cohort stratified into Low, Medium, and High iTRAC levels. G. Hazard Ratio (HR) with a 95% confidence interval (CI) for the risk of metastasis in each iTRAC group relative to the Low group for the chemotherapy-treated panSTS cohort. P-values for each HR are shown above each condition. The dashed horizontal line indicates an HR of 1.

### Chemotherapy is associated with a decreased MFS in Low iTRAC patients and an increased MFS in Medium iTRAC Patients

Using WGS and leimoyosarcoma cohort, we have previously shown that in Low iTRAC group, patients treated with chemotherapy have increased metastatic risk compared to patients not treated (Benhaddou *et al*. 2021). To check whether this observation holds in panSTS cohort using RNAseq, we split panSTS patients having chemotherapy (59 patients) into Low, Medium and High levels using the thresholds determined with patients not treated with chemotherapy. First we observed that the prognostic value of iTRAC stratification (Low, Medium and High) changed significantly (fig 1C) with the poorer MFS for the Low iTRAC group (HR = 4.95; P = 2.22×10-3; fig 1D), the better for the High iTRAC group and an intermediate MFS for the Medium group. Thus, we analyzed separately each iTRAC group. In Low iTRAC group, patients who received chemotherapy have a higher metastatic risk than those who did not (fig 1E). Interestingly, in the Medium iTRAC group, patients who received chemotherapy have a better MFS (fig 1F, HR=3.01, P = 3.33 10-2). Finally, no significant difference in MFS was observed between patients with or without chemotherapy in High iTRAC (fig 1G). Therefore, Low and Medium levels of iTRAC for patients treated with chemotherapy are associated with different MFS contrary to patients not treated with chemotherapy. Taken together, these results show that Low iTRAC patients treated with chemotherapy have a poorer prognosis whereas Medium iTRAC patients treated with chemotherapy have a better prognosis than those not treated with chemotherapy.

### iTRAC performs better than FNCLCC and CINSARC in stratifying metastatic risk in panSTS cohort

The current standard in sarcomas for predicting patient evolution is the histological FNCLCC grading system (Coindre *et al*. 2001). Complexity INdex in SARComa (CINSARC), which is the main expression-based prognostic signature of sarcomas, is currently under prospective clinical investigation for patients stratification (Chibon *et al*. 2010; Filleron *et al*. 2020). FNCLCC grading did not significantly split the panSTS cohort into groups with different metastatic evolution for both chemotherapy-treated and not treated patients (fig 2A and 2B). Interestingly, for CINSARC, while there is a significant difference in MFS between groups C1 and C2 for patients without chemotherapy, there is no statistically different MFS between C1 and C2 for patients with chemotherapy (fig 2C and 2D). Consequently, no significant difference in MFS between patients treated or not by chemotherapy was observed in each group of FNCLCC grade and CINSARC (fig 3A, 3C, and fig 3B, 3D). Multivariate analysis with a Cox proportional hazard model comparing iTRAC, CINSARC and FNCLCC grading shows (fig 4) that in patients who did not receive chemotherapy, Medium iTRAC is significantly associated (P<0.001) with increased HR of metastatic risk but not FNCLCC nor CINSARC (fig 4A). Conversely, in chemotherapy-treated patients, Low iTRAC, but not FNCLCC or CINSARC, was significantly associated with increased metastatic risk compared to High iTRAC, while the Medium iTRAC group was no longer significantly associated with increased MFS (fig 4B). Taken together, these results show that iTRAC outperforms both FNCLCC grading system and CINSARC signature in stratifying metastatic risk both in chemotherapy treated and not treated patients.

**Figure 2:**
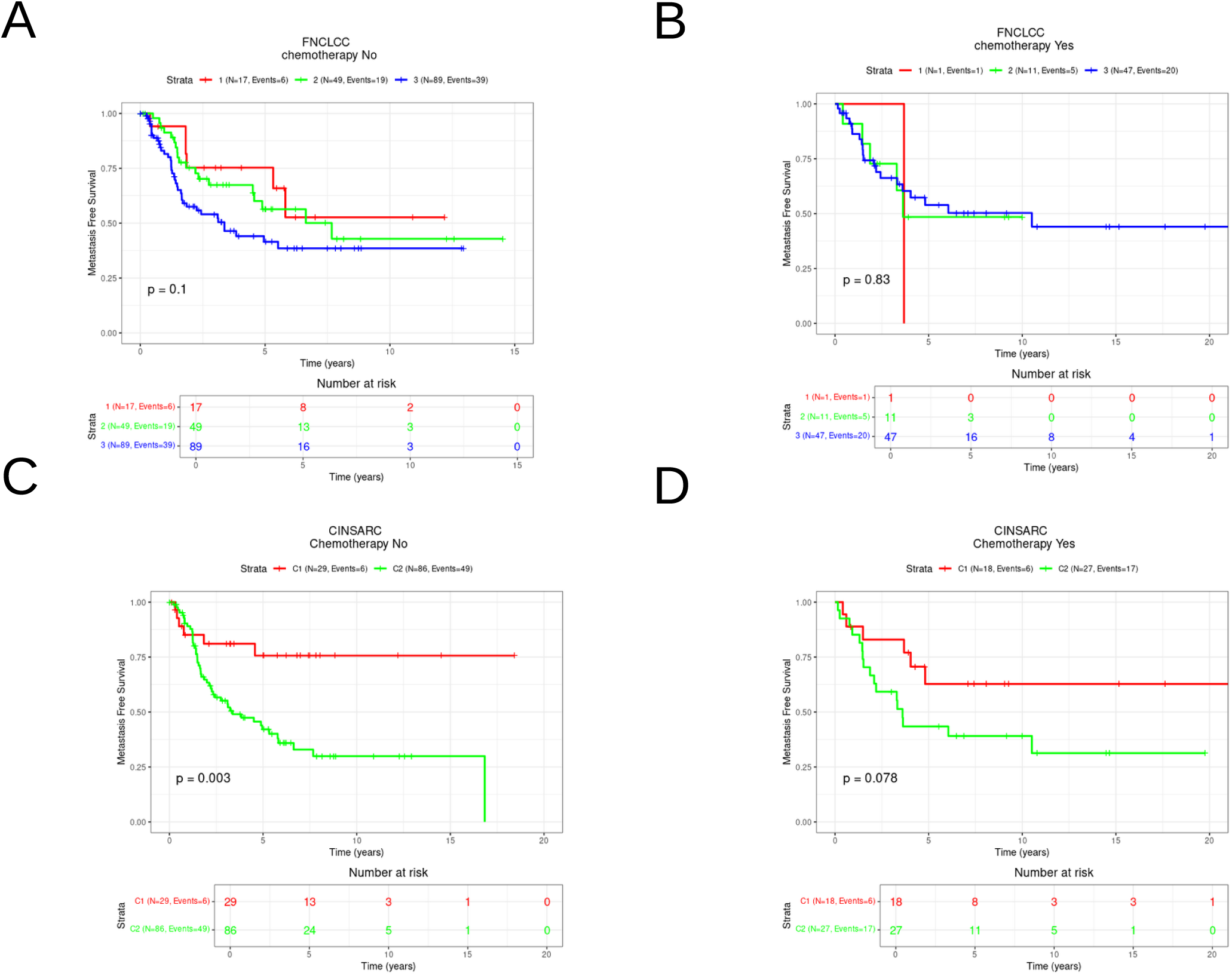
MFS stratification of chemotherapy-treated and not treated patients using FNCLCC and CINSARC. A. Kaplan-Meier plot for patients without chemotherapy stratified by FNCLCC. B. Kaplan-Meier plot for patients treated with chemotherapy stratified by FNCLCC. C Kaplan-Meier plot for patients without chemotherapy stratified by CINSARC A Kaplan-Meier plot for patients treated with chemotherapy stratified by CINSARC.

**Figure 3.**
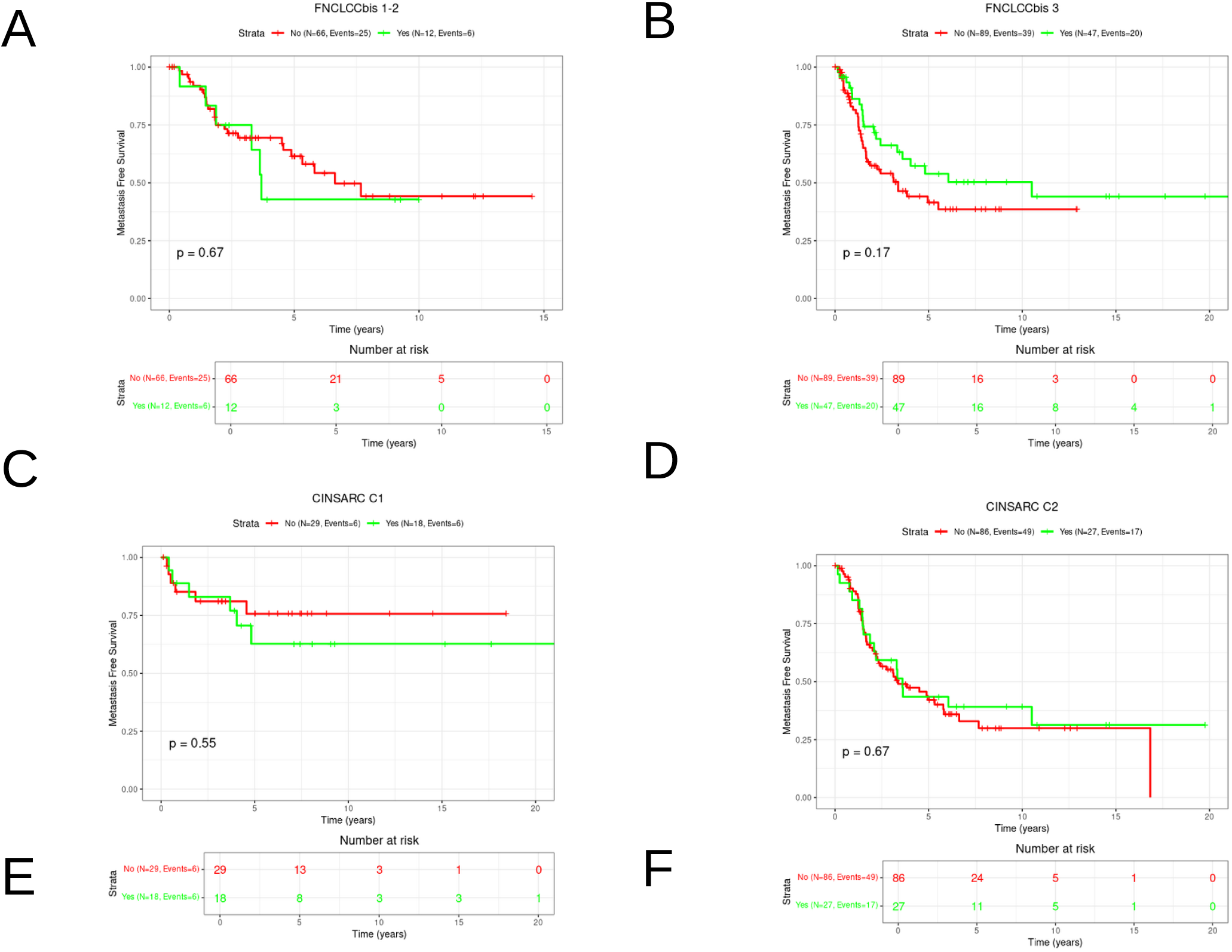
FNCLCC and CINSARC levels have similar MFS in between treated and not treated patients. A,B, Kaplan-Meier for patient with or without chemotherapy in FNCLCC grades 1-2, and 3 respectively. B,D, Kaplan-Meier for patient with or without chemotherapy in CINSARC groups C1, and C2,

**Figure 4.**
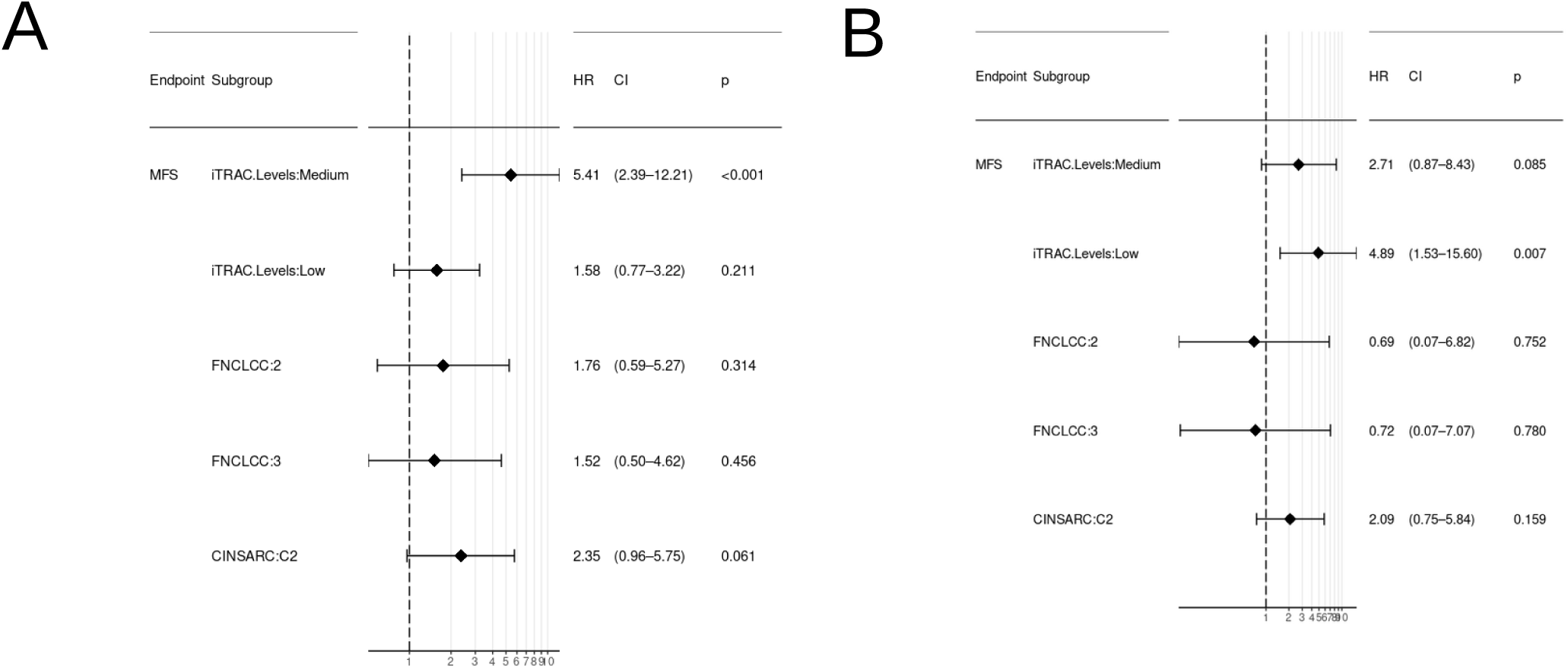
MFS analysis using Multivariate cox regression using iTRAC FNCLCC and CINSARC as covariates in A. in PanSTS patients not treated with chemotherapy B. Treated with chemotherapy.

## Discussion

### Medium levels of iTRAC measured on RNAseq is associated with poor prognosis

We have previously shown that Medium level of iTRAC in LMS, measured on WGS, is associated with poorer clinical outcome than High and Low levels (Benhaddou *et al*. 2021). Here we established the same conclusion by measuring iTRAC by RNAseq in a cohort of 226 pleomorphic STS. This finding opens the door to a wide prospective clinical evaluation of MAGIC in the clinic for these pleomorphic sarcomas and other cancers for which the non-linear relationship between GIN and clinical outcome was described, as for in ER^-^ breast cancers, lung, ovarian and gastric cancers (Roylance *et al*. 2011; Birkbak *et al*. 2011).

### Medium level of iTRAC patients have a better prognosis in treated than in non-treated patients

Our findings reveal that STS patients with Medium iTRAC exhibit distinct prognosis based on whether they received chemotherapy treatment or not. This observation adds a novel dimension to our understanding of the interplay between GIN and chemotherapy efficacy.

Previous studies have established a link between GIN and poor prognosis in solid tumors (Carter *et al*. 2006; Walther, Houlston and Tomlinson 2008; Birkbak *et al*. 2011). However, there is compelling evidence suggesting that certain cancer treatments, which induce GIN, may be more effective in cells with a higher baseline rate of aneuploidy. For instance, it has been shown that the efficacy of treatments like Paclitaxel and radiation therapy improves under higher levels of GIN (Bakhoum *et al*. 2015), (Janssen, Kops, et Medema 2009), and (Zasadil *et al*. 2014). Similarly, rectal adenocarcinoma patients with elevated pre-treatment aneuploidy are more likely to respond positively to chemoradiation therapy (Zaki *et al*. 2014).

Several chemotherapeutic drugs have been shown to induce GIN *in vitro*, leading to additional structural variations (SVs) that surpass a threshold incompatible with cell survival (Andor *et al*. 2016; Kim *et al*. 2016; Bakhoum *et al*. 2015). This mechanism may underpin the improved prognosis observed in treated patients with medium iTRAC, suggesting that these treatments leverage the existing GIN to drive cancer cells beyond survivable limits.

Interestingly, Cahill and colleagues proposed that there is a need for an optimal level of GIN for tumor aggressiveness. Beyond this optimal level, additional instability generates non-viable karyotypes and provides no competitive advantage to cancer cells (Cahill *et al*. 1999). This concept is crucial for understanding why patients with medium levels of iTRAC likely benefit the most from chemotherapy.

In contrast, untreated patients with medium iTRAC might experience the detrimental effects of medium levels GIN without the benefit of systemic therapeutic intervention, resulting in a poorer prognosis. This dichotomy underscores the importance of treatment in managing medium level GIN and highlights the potential for personalized therapeutic strategies based on iTRAC levels. Our results, therefore, provide new insights supporting the notion that GIN, while generally associated with poor outcomes, can be exploited therapeutically to improve patient prognosis in specific contexts.

### High level of iTRAC is not associated with a different MFS in treated or not treated patients

Our findings indicate that high level of iTRAC is not associated with a significant difference in metastasis-free survival (MFS) between patients treated with chemotherapy and those who were not. This result aligns with previous research on GIN and cancer outcomes.

Cahill and colleagues (Cahill *et al*. 1999) proposed that GIN contributes to oncogenesis only up to a certain threshold. Beyond this threshold, the level of instability is likely to produce cells with unviable karyotypes, which are not sustainable for tumor progression. This concept is also crucial in understanding why high levels of iTRAC do not confer additional survival benefits in the context of chemotherapy. In fact, when GIN surpasses a certain point, it may result in excessive cellular dysfunction, to which cancer cells became resistant by negative pressure selection, rendering further therapeutic interventions less effective.

The lack of prognostic difference in High iTRAC patients treated vs untreated is in line with what was reported by Birkbak and colleagues showing that the improved outcomes observed in tumors with very high aneuploidy appear to be independent of treatment (Birkbak *et al*. 2011). This suggests that once a high level of GIN is reached, the biological behavior of the tumor is driven more by the intrinsic instability rather than the effects of chemotherapy.

### Low level of iTRAC is associated with an increase in MFS for patients treated with chemotherapy

Chemotherapy increases survival in patients in several cancers (Von Minckwitz *et al*. 2012; Rossi *et al*. 2015) but still remains controversial in STS. There are many patients who do not benefit from chemotherapy and few studies report that in some cases it might participate to metastatic relapse (Karagiannis, Condeelis, et Oktay 2018) by inducing cytokines (Daenen *et al*. 2014) and proinflammatory circuits (Byrd-Leifer *et al*. 2001). Interestingly, Low GIN cancer cells contain mainly stable genome (Birkbak *et al*. 2011). Thus they are the closest to a normal cell and this may explain at least in part their good prognosis. This may also be the case for Low iTRAC patients. The fact that in Low iTRAC group, patients having chemotherapy have decreased MFS compared to patients not treated, may be due to a global increase of GIN so that they become of medium level which is associated with more aggressive phenotype, either by inducing individual chromosomal instability or by targeting preferentially less rearranged and fast proliferating cells in an heterogenous cellular population.

Our findings highlight the nuanced role of GIN in cancer treatment outcomes. Low iTRAC levels are associated with increased MFS in untreated patients, while Medium iTRAC patients benefit from chemotherapy, suggesting an optimal window of instability for therapeutic exploitation. In contrast, high iTRAC levels do not show significant treatment benefits, likely due to the deleterious effects of resistance of cancer cells to excessive additional GIN on cancer cells fitness. These insights underscore the importance of personalized therapeutic strategies that consider a patient’s specific GIN profile to maximize treatment efficacy and improve prognosis.

Several confounding factors, like tumor grade and histotype, may undermine this hypothesis. We can’t rule out the possibility that distinct prognosis in treated and non-treated patients could also be due to bias in patients selection inherent to retrospective studies. Therefore, it is only a controlled prospective analysis that will give the final answer to this question. Finally, we do not exclude the possibility that the Low iTRAC patients have different immune infiltrate than Medium or High ones and which portends them to be of higher risk of metastasis upon chemotherapy. Further controlled prospective studies are needed to explore the relationship between the different levels of iTRAC, immune infiltrate/tumor microenvironnement and chemotherapeutic response.

### MAGIC, from whole genome to RNAseq

MAGIC was originally developed using BP of structural variations (SVs) identified through Whole Genome Sequencing, including insertions, deletions and translocations. Both iTRAC and iRACIN significantly stratified patients and demonstrated a non-linear relationship with clinical outcomes (Benhaddou *et al*. 2021). Notably, only iTRAC showed relevance in stratifying metastatic risk for chemotherapy treated and non-treated patients. Here, we applied MAGIC to RNA sequencing data using BP of fusion transcripts. Fusion transcripts arise from various events such as insertions, deletions and translocations (Aguilera et Gómez-González 2008). Notably, only iTRAC was measurable in RNA-seq data underscoring its unique capability to capture transcription-associated GIN. Therefore, the application of iTRAC to RNA-seq data effectively captures the complexity and relevance of transcription-associated GIN measured through WGS. This specificity makes iTRAC particularly relevant for identifying functionally significant genomic regions that directly impact gene expression and tumor behavior, providing valuable prognostic and therapeutic insights. Furthermore, fusion transcripts have prognostic relevance. For example, in breast cancer, associations have been reported between the number of fusion transcripts, copy number events, gene expression profiles and clinical outcomes (Thompson *et al*. 2016). Additionally, high frequencies of fusion transcripts detected in breast cancer patients correlate with poor outcomes (Ma *et al*. 2014).

In summary, MAGIC’s adaptation for RNA sequencing within the iTRAC biomarker distills the broader concept of GIN into a focused analysis of fusion transcripts. This approach enhances our ability to infer transcription-associated GIN and its implication for cancer prognosis and treatment strategies.

### From quantity to enrichment: iTRAC and the qualitative dimension of genomic instability

A key nuance in the interpretation of Hscore lies in distinguishing between quantitative measures of genomic instability and qualitative enrichment in specific genomic regions. While the total number of BP reflects the overall burden of structural variations, Hscore captures the localized "hotspotness" of BPs, providing insights into the mechanistic underpinnings of GIN. Interestingly, using iPART, stratification of panSTS cohort by total BP counts is less effective than by number of BP in iTRAC (supplementary figure 1), further stressing the importance of regional context in genomic instability. This suggests that not all BP contribute equally to tumor behavior or treatment response. Those occurring within iTRAC-defined regions may reflect functionally relevant alterations, such as replication stress or fragile site collapse, thereby offering superior predictive and prognostic values compared to global breakpoint burden alone. Furthermore, Low Hscore may reflect a diffuse or untargeted instability, potentially linked to stochastic processes or generalized cellular constraints. In contrast, high Hscore indicates specific mechanistic processes such as transcription-replication collisions or transcription-associated stress. The intermediate Hscore likely reflects a convergence of transcription-associated GIN and cellular constraints such as clonal selection, leading to a state that is both permissive to genomic diversity and conducive to aggressive tumor behavior. These insights have direct clinical implications, as patients with high transcriptional stress (High iTRAC) may benefit from therapies targeting transcription-associated mechanisms, such as PARP inhibitors, while patients with low or diffuse instability might require alternative strategies. Indeed, Petropoulos *et al*. 2024 elegantly demonstrated that Transcription– replication conflicts underlie sensitivity to PARP inhibitors (Petropoulos *et al*. 2024). Future studies should aim to further elucidate the biological drivers of Hscore enrichment and validate its utility as a biomarker for precision oncology. This work underscores the need to move beyond bulk measures of mutation load and instead focusing on the structural and functional contexts of genomic instability.

## Data Availability

All data produced in the present work are contained in the manuscript

## Acknowledgments

This work was supported by Région Occitanie, INCa (Institut National du Cancer), Inserm, Inserm Transfert, IUCT/ICR (Institut universitaire de cancerologie de Toulouse/Institut Claudius Regaud). We thank these institutions for their financial and strategic support of this research.

## Ethics declaration

The samples used in this study are part of the Biological Resources Center of Bergonie Cancer Institute (CRB-IB). Accordance with the French Public Health Code (articles L. 1243-4 and R. 1243-61), the CRB-IB has received the agreement from the French authorities to delivered samples for scientific research (number AC-2008-812). The samples come from care and are re-qualified for research as part of the ICGC program (International Cancer Genome Consortium), with patient consent. The project was approved by the Bergonie Institute ethic committee (scientific advisory board).

**supplementary Figure 1:**
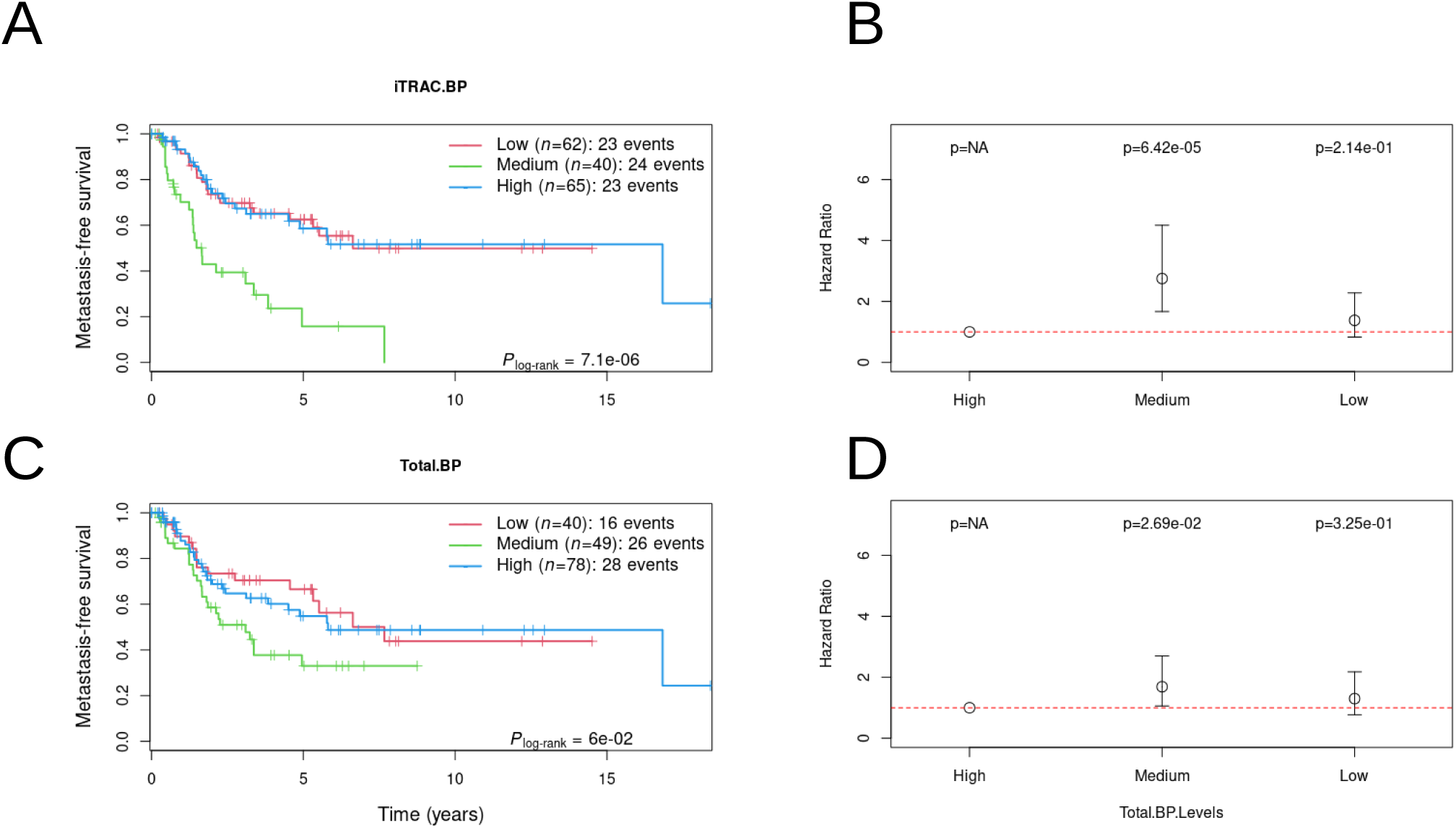
Stratification of panDF cohort using iTRAC BP counts is more significant than using total BP count : A,B Kaplan Meier and Hazard ratio for stratification of panSTS cohort using iTRAC BP counts. CD, aplan Meier and Hazard ratio for stratification of panSTS cohort using total BP counts.

**Supplemental table 1:** STS patients characteristics.

| Characteristics | panSTS cohort (n=226) |
| --- | --- |
| Median follow-up (months)[IQR] | 2.35[4.15] |
| Median age at diagnosis (years)[IQR] | 64[17.75] |
| Female sex ( %) | 59.93(26.5) |
| FNCLCC grade ( %) |  |
| 1 and 2 | 78(34.5) |
| 3 | 136(60.2) |
| ND | 12(5.3) |
| CINSARC ( %) |  |
| C1 | 43(20.8) |
| C2 | 113(50) |
| ND | 66(29.2) |
| Histological type ( %) |  |
| Leimyosarcomas | 122(54) |
| Undifferentiated pleomorphic sarcoma | 45(19.9) |
| Liposarcoma - dedifferentiated | 21(9.3) |
| Myxofibrosarcoma | 18(8) |
| Liposarcoma - pleomorphic | 8(3.5) |
| Pleomorphic rhabdomyosarcoma | 5(2.2) |
| Endometrial stromal sarcoma | 3(1.3) |
| Low grade fibromyxoid sarcoma | 2(0.9) |
| Malignant solitary fibrous tumor | 2(0.9) |
| Metastatic events(%) |  |
| Yes | 96(42.48) |
| No | 130(57.52) |
| Therapeutic management(%) |  |
| chemotherapy | 59(26) |
| radiotherapy | 113(50) |
| Radiotherapy lines (% radiotherapy) | 57(22.62) |
| First | 1(0.8) |
| Second | 87(77) |
| Third | 25(22.2) |

ND : Not Determined
IQR : InterQuartile Range

